# Foundational research and NIH funding enabling Emergency Use Authorization of remdesivir for COVID-19

**DOI:** 10.1101/2020.07.01.20144576

**Authors:** Ekaterina Galkina Cleary, Matthew J. Jackson, Zoë Folchman-Wagner, Fred D. Ledley

## Abstract

Emergency Use Authorization for remdesivir months after discovery of COVID-19 is unprecedented. Typically, decades of research and public-sector funding are required to establish the mature body of foundational research requisite for efficient, targeted drug discovery and development. This work quantifies the body of research related to remdesivir’s biological target, RNA-dependent RNA polymerase (RdRp), or parent chemical structure, nucleoside analogs (NcAn), through 2019, as well as NIH funding for this research 2000–2019. There were 6,567 RdRp-related publications in PubMed, including 1,263 with NIH support, and 11,073 NcAn-related publications, including 2,319 with NIH support. NIH support for RdRp research comprised 2,203 Project Years with Costs of $1,875 million. NIH support for NcAn research comprised 4,607 Project Years with Costs of $4,612 million. Research Project grants accounted for 63% and 48% of Project Years for RdRp and NcAn respectively, but only 19% and 12% of Project Costs. Analytical modeling of research maturation estimates that RdRp and NcAn research passed an established maturity threshold in 2008 and 1994 respectively. Of 97 investigational compounds targeting RdRp since 1989, the three authorized for use entered clinical trials after both thresholds. This work demonstrates the scale of foundational research on the biological target and parent chemical structure of remdesivir that supported its discovery and development for COVID-19. This work identifies $6.5 billion in NIH funding for research leading to remdesivir, underscoring the role of public sector investments in basic research and research infrastructure that underlie new drugs and the response to emergent disease.

**SIGNIFICANCE STATEMENT:** Emergency Use Authorization of remdesivir for treating COVID-19 four months after discovery of this virus was enabled by decades of research on the drug’s biological target as well as other medicines with related chemical structures. The NIH contributed 6,800 years of grant funding to this research, totaling $6.5 billion (2000–2019), including funding for both investigator-initiated research and research infrastructure. Of this, $46.5 million was for research directly related to remdesivir. This analysis demonstrates the importance of a robust body of foundational research in responding rapidly to emergent diseases, and the substantial NIH contribution to this research. It also underscores the scale and significance of the public-sector investments that enable new drug discovery and development.

---

The Emergency Use Authorization of remdesivir for treating SARS-CoV-2 (COVID-19)(1) four months after the discovery of this pathogen is unprecedented. Typically, decades are required to translate research evidence into clinical practice or approved products.(2) For example, a study characterizing the research and development timelines of 113 first-in-class drugs approved from 1999–2013 described a median 20-year lag between the initial identification of a drug target, pathway, or chemotype, and first drug approval.(3) Similarly, studies with an analytical model of research maturation estimated the time from initiation of research related to a novel drug target to first approval of a drug against that target was more than 30 years.(4, 5) While efforts to reduce this translational lag commonly focus on the length of time required for clinical trials and regulatory review,(6) evidence suggests the longer, rate-limiting step is the time required for the basic research required to establish the requisite scientific foundation for successful development.(3-5, 7, 8)

Basic research is funded primarily by the public sector.(9) Previous studies have shown that NIH funding contributed to published research underlying every new drug approved from 2010–2016, with >90% of this funding representing basic research on the biological target, rather than applied research on the drugs themselves.(10, 11) These studies also show that this research often involves substantial spillover effects, in which research performed in various therapeutic areas contributes to the scientific foundation for products in another area.(12)

Research on strategic product development in different fields of technology has identified predictable patterns in the advance of science or technology. These studies also demonstrate the enabling role for a mature foundation of research and technology readiness in successful product development.(13, 14) The same pattern is evident in biomedical research and targeted drug development. Analytical modeling demonstrates that few targeted therapeutics are successfully developed until foundational research on both the biological target and the nature of the molecular entity passes a defined established point.(4, 5, 8, 15)

This work examines the body of research on remdesivir’s biological target, RNA-dependent RNA polymerase (RdRp), and its parent chemical structure, nucleoside analogs (NcAn). Specifically, we quantify the number of research publications in these two areas, model the advance of this research and thresholds of maturation, describe the number of investigational products targeted to RdRp and timelines of development for the three RdRp inhibitors currently in clinical use, and detail the NIH investments made in this research. Collectively, these data describe the foundational research, and NIH investment in this research, that enabled rapid development of remdesivir in response to the COVID-19 pandemic.

These results are considered in the context of intense societal debate over the potential pricing of remdesivir by its manufacturer, Gilead Sciences.(16-18) One aspect of this debate concerns investments made by the public sector in the early development of this drug and its subsequent application to COVID-19.(19, 20) This work provides an estimate of the NIH contribution to remdesivir through the end of 2019 by considering the foundational research that enabled the discovery and development of this product as well as the NIH contribution to this work.

## RESULTS

### Publications on Remdesivir’s Target and Chemical Type

PubMed searches using optimized search terms (Supplemental Table 1) identified 6,567 PubMed Identifiers (PMID) related to RdRp and 11,073 PMID related to NcAn (Table 1). Research on RdRp originated in the 1950s, grew exponentially from the 1980s, and has been level from 2011 to the present (Figure 1A). Research on NcAn also dates from the 1950s, increased exponentially beginning in the 1980s in conjunction with the emergence of HIV, and has been decreasing slightly from the late 1990s to the present (Figure 1B).

**Table 1.**
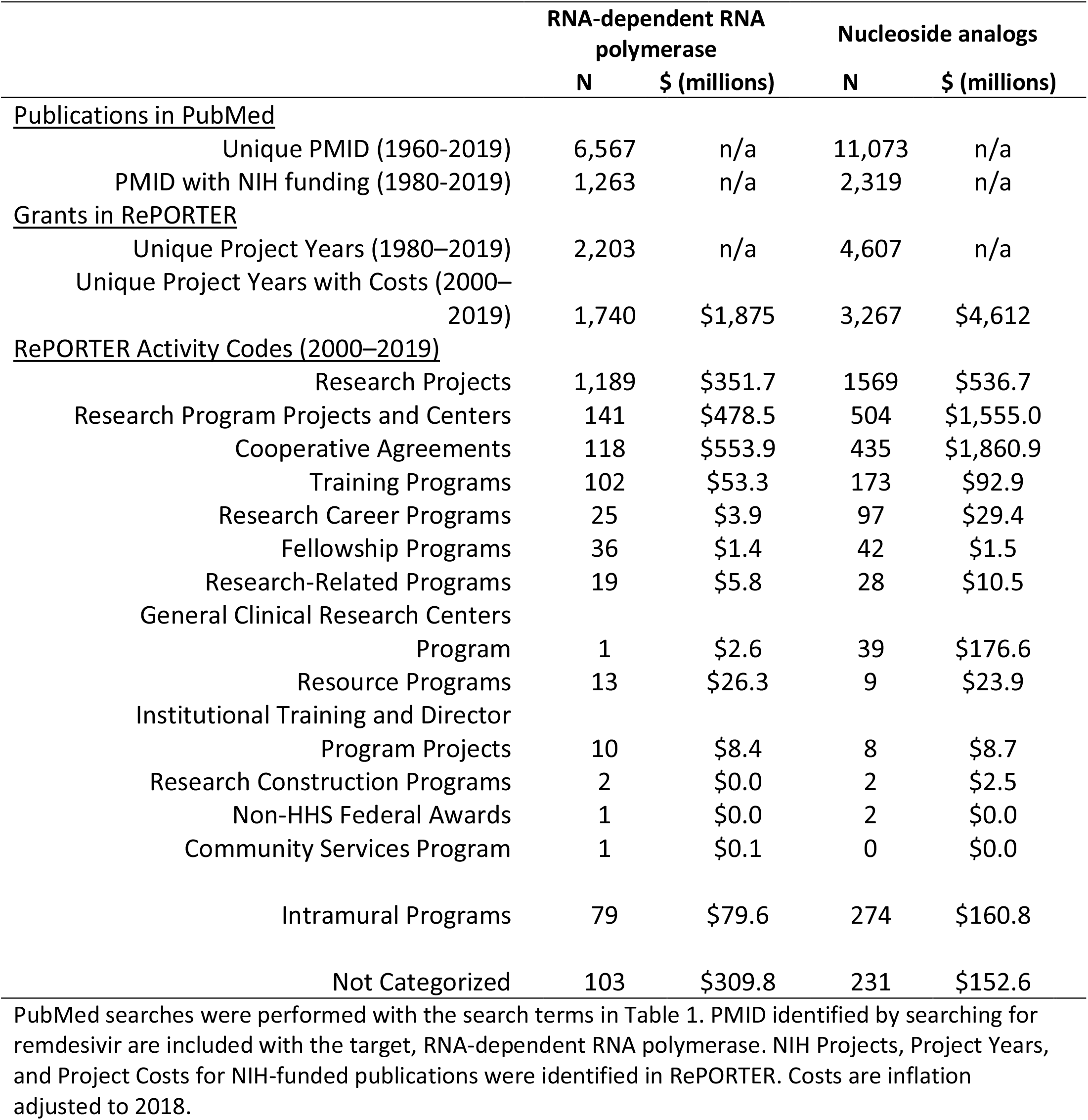
**Publications, Project Years of NIH funding, and Projects Costs related to research on remdesivir, RNA-dependent RNA polymerase, and nucleoside analogs**.

**Figure 1.**
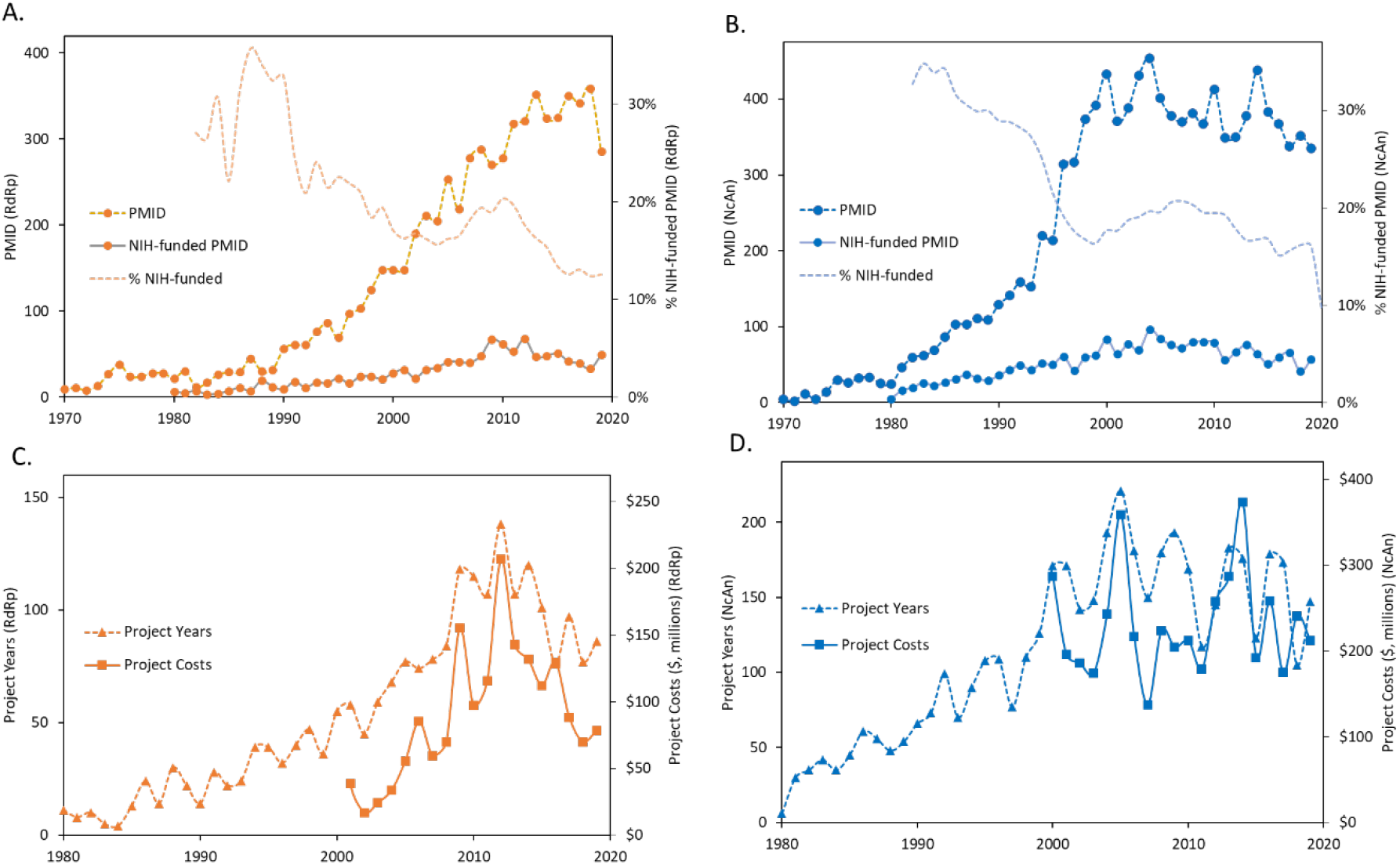
Time course of publications (PMID), NIH-funded Project Years, and Project Costs related to research on remdesivir, RNA-dependent RNA polymerase (RdRp), or nucleoside analogs (NcAn). (A) PMID and NIH-funded PMID (left axis) related to RdRp and percentage of PMID supported by NIH funding; (B) PMID and NIH-funded PMID (left axis) related to NcAn and by NIH funding (right axis); (C) NIH Project Years and Costs related to RdRp; (D) NIH Project Years and Costs related to NcAn. PubMed searches were performed with the search terms in Table 1. PMID identified by searching for remdesivir are included with the target, RNA-dependent RNA polymerase. NIH Projects, Project Years, and Project Costs for NIH-funded publications were identified in RePORTER. Costs are inflation adjusted to 2018.

### NIH Funding for Published Research

NIH funding was associated with 1,263 (19%) of publications on RdRp and 2,319 (21%) of publications on NcAn (Table 1). For both RdRp and NcAn, the fraction of publications with NIH support decreased steadily from >30% prior to 1990 to 13–16% after 2015 (Figure 1A, 1B).

NIH support for RdRp research comprised 2,203 Project Years of funding totaling $1,875 million (Table 1, Figure 1C). While Research Projects represented the largest fraction of Project Years since 2000 (63%) and had Costs of $351.7 million, Research Projects represented only 19% of total Costs, less than the $478 million in Research Program Projects (26%) or $553.9 million in Cooperative Agreements (30%) (Table 1). NIH support for RdRp research increased steadily from the 1980s, and decreased from 2012–2019 (Figure 1D).

NIH support for NcAn research comprised 4,607 Project Years of funding totaling $4.6 billion (Table 1). While Research Projects represented the largest fraction of Project Years (48%) and had Costs of $537 million, Research Projects represented only 12% of total Costs, less than the $1.56 billion for Research Program Projects and Centers (34%) or $1.9 billion in Cooperative Agreements (40%) (Table 1). NIH support for NcAn research increased up to 2000 and remained stable through 2019 (Figure 1D).

### Topic Focus of NIH-Funded RdRp and NcAn Research

The large majority of NIH-funded publications on RdRp (89%) mentioned the term virus or viral, with the majority accounted for by research related to Hepatitis, influenza, Coronavirus, Zika, and Ebola virus (Figure 2A), while 19% contained the wild card term “immun*” and 1.3% mentioned cancer. The timeline of research involving various RNA viruses can be viewed through an interactive graphic online. *(Note to Editor: hyperlink to: https://bit.ly/RdRpRemdesivir)*

**Figure 2.**
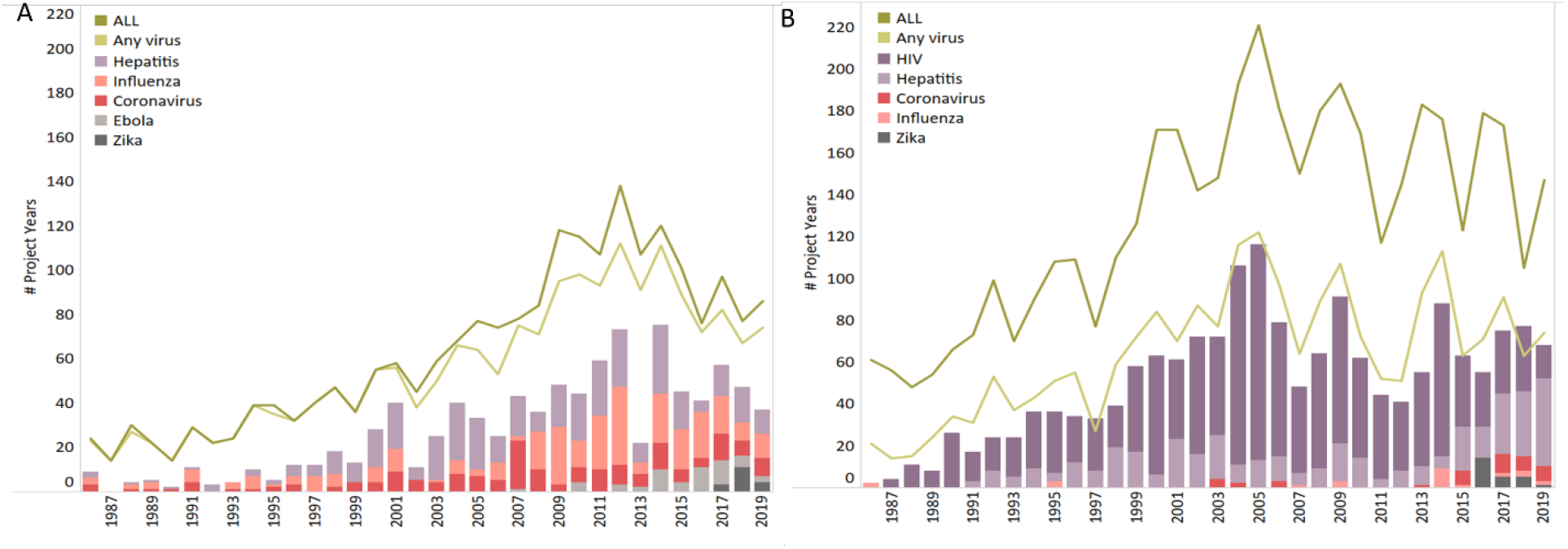
Topic focus of NIH-funded Project Years related to RNA-dependent RNA polymerase (RdRp) or nucleotide analogs (NcAn) over time as determined by text analysis of abstracts from NIH-funded PMID. Figure shows the total number of Project Years along with the number associated with publications mentioning any virus (lines), and the number associated with publications mentioning hepatitis, HIV, influenza, Coronavirus, Ebola, or Zika viruses (stacked bars). (A) Project Years related to RdRp; (B) Project Years associated with NcAn.

Of the NIH-funded publications related to NcAn, only 49% mentioned the term virus or viral (Figure 2B), 28% contained the wild card term “immun*” and 15% were related to cancer. The most commonly mentioned viruses were HIV, Hepatitis, Coronavirus, influenza, and Zika virus, respectively. Funding for influenza and Coronavirus was sporadic. Funding for Zika research rose following the 2015-16 outbreak.

### Maturation of RdRp and NcAn Research and Timing of Drug Approvals

The Technology Innovation Maturation Evaluation (TIME) model quantifies the growth of a corpus of published literature as well as inflection points related to research maturation. The calculated *initiation* point, or point of maximum acceleration of publication activity, corresponds to the time of early discoveries that lead to exponential growth in research activity. The calculated *established* point, or point of maximum slowing of publication activity, corresponds to a time before which targeted therapeutics are successfully developed.(4, 8) Plotted on a log scale with cumulative publications, this function exhibits the logistic “S-curve” characteristic of technology maturation models in other fields.(13)

Figure 3A and Figure 3B show annual publications and TIME model curve fit for RdRp and NcAn respectively. Figures 3C and 3D show cumulative publications and the TIME model curve plotted on log scale. In this projection, it is evident that RdRp research progressed through two successive S curves. The first has a calculated *initiation* point (Ti) of 1962 and exhibited exponential growth through the late 1960s. The second exhibited exponential growth through the 1990s and early 2000s, reaching a calculated *established* point (Te) of 2008. This pattern of successive S-curves is consistent with evidence from other fields that technological progress classically proceeds through a series of novel innovations, each of which introduces a new period of exponential advance leading to a limit.(13, 14)

**Figure 3.**
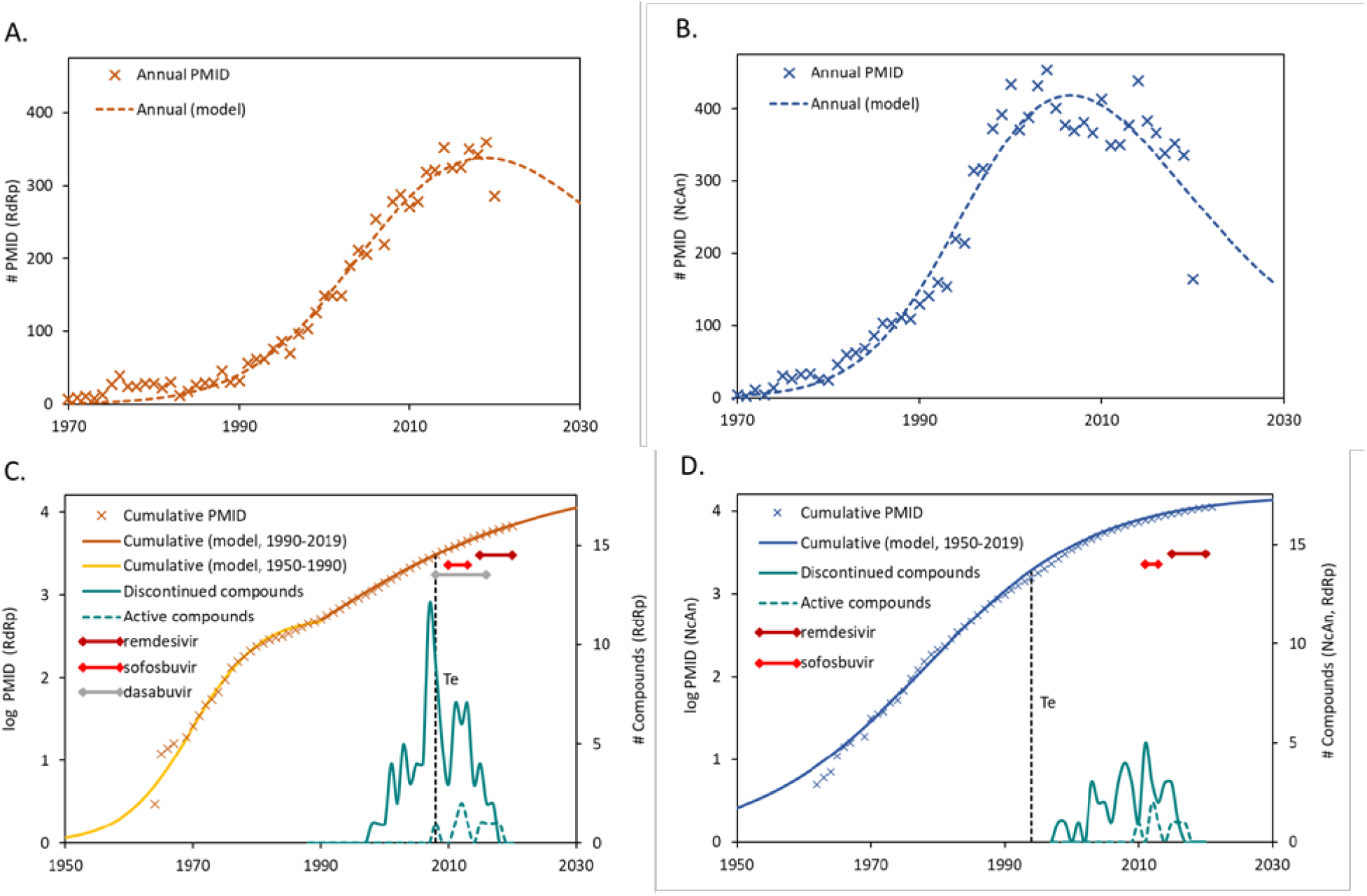
Timeline of research maturation and investigational compounds targeted to RNA-dependent RNA polymerase (RdRp) with a nucleoside analog (NcAn) parent chemical structure. The Technology Innovation Maturation Evaluation (TIME) model was used to estimate the *established* point for research on RdRp and NcAn. (A) Annual PMID related to RdRp and TIME model approximation; (B) Annual PMID related to NcAn and TIME model approximation; (C) Cumulative PMID related to RdRp (left axis) along with number of compounds targeted to RdRp entering development over time (right axis), including discontinued compounds (solid line) and compounds still in active development (dashed line). Also shown are the timelines from the initiation of clinical trials to approval for clinical use for remdesivir, sofosbuvir, and dasabuvir; (D) Cumulative PMID related to NcAn (left axis) along with number of NcAn compounds targeted to RdRp entering development over time (right axis), including discontinued compounds (solid line) and compounds still in active development (dashed line). Also shown are the timelines from the initiation of clinical trials to approval for clinical use for remdesivir and sofosbuvir.

PharmaProjects lists 97 investigational drugs targeted against RdRp since 1989. Figure 3C shows the clinical development timelines for the two approved products, sofosbuvir and dasabuvir, as well as remdesivir. Also shown is the distribution of first reported development for 94 drugs that have been discontinued or remain in clinical development. These data show that each of the products now in clinical use entered clinical trials only after RdRp technology reached the *established* point and were approved 5, 8, and 12 years after this point (Figure 3C).

Of the 97 investigational drugs targeted against RdRp, 47 are NcAn. This includes two in clinical use, sofosbuvir and remdesivir, 40 discontinued, and five in development at the end of 2019. Of the 50 that are not NcAn, one is approved, dasabuvir, and three remain in development, including silibinin sodium hemisuccinate (approved in the EU) as well as CC-31244 and cephaeline, which are reported as being in development at the end of 2019 by PharmaProjects.

Research on NcAn followed a single S-curve with calculated initiation points of 1961 and a calculated *e*s*tablished* point of 1994 (Figure 3D). More than 60 NcAn drugs and combinations are approved by the FDA for cancer or various viral diseases including HIV, hepatitis-C (HCV), or herpes simplex virus.(21-24) Both sofosbuvir and remdesivir entered clinical trials long after the *established* point of NcAn research and entered clinical practice 19 and 26 years after this point (Figure 3D).

### Research and Development of Remdesivir Through 2019

A PubMed search for remdesivir (GS-5734) identified 23 published before the end of 2019, of which six had NIH support indicated in RePORTER. There were an additional 106 PMID from January through May 12, 2020, but RePORTER data on NIH involvement in this research is not yet available.

The six papers published through 2019 were supported by 18 Project Years of NIH funding totaling $46.52 million in Project Costs from 2000-2019 (Supplemental Table 2). This result is similar to the total described in a recent KEI Briefing Note detailing the role of the federal government in the development of remdesivir (20). The KEI Briefing Note tracked specific events in product development from 2014 through May 2020, identifying seven NIH Projects mentioned in the acknowledgements and having Project Costs in RePORTER (Supplemental Table 2). The methods used in the KEI Briefing note and the present work differ substantially. The KEI Briefing Note included total Project Costs of each NIH Project acknowledged in these publications, but explicitly excluded the Costs of Program Projects and Centers or Projects related to training. The present work considers only the Project Year corresponding to the specific publication date, and does include Costs of Program Projects, Centers, and Training Projects.

Supplemental Table 3 details non-NIH sources of funding acknowledged in the 23 publications on remdesivir published through 2019 as well as co-authorship from these institutions. Of the 15 publications acknowledging sources of support, the majority reported involvement by Gilead Sciences. Other sources of funding most commonly cited were the Center for Disease Control and agencies within the Department of Defense. Project Costs for non-NIH sources of funding are not publicly available.

## DISCUSSION

This study examined the body of foundational research underlying remdesivir in the context of studies suggesting that efficient development of targeted therapeutics is critically dependent on a mature body of basic and applied research.(4, 5, 25, 26) Quantitative analysis demonstrates that decades of research on RdRp and NcAn generated established bodies of research on both the drug target and parent chemical structure of remdesivir at least ten years before the emergence of COVID-19 and the subsequent Emergency Use Authorization of remdesivir. This analysis also demonstrates that the NIH contributed over $6 billion to this body of research.

The quantitative picture of research on RdRp and NcAn described here closely parallels the history of these fields. The calculated *initiation* point for research on RdRp corresponds to the recognition of RNA-dependent RNA polymerase activity in viruses such as poliovirus, mengovirus, and Newcastle disease virus in the 1960s.(27-29) This research introduced a period of exponential advance and generation of an extensive body of literature on the structure and function of homologous RdRp enzymes from various positive and negative stranded RNA viruses.(30) Text analysis demonstrates that a large fraction of this research has involved endemic viruses (e.g., HCV) or emerging epidemic viruses including Ebola and Coronavirus.

The first NcAn drugs arose from the discovery of naturally occurring products comprising nucleobases with anticancer activity(31). Early studies on NcAn compounds demonstrated that these compounds had poor pharmaceutical properties, and decades of research focused on developing strategies to increase the lipophilicity and bioavailability of the molecules and facilitate metabolism of the prodrug to the active triphosphate.(32, 33) Today, NcAn drugs are commonly inactive prodrugs with suitable pharmaceutical properties, which must be metabolized to the biologically active compound once absorbed into the body.

The development of remdesivir benefited directly from both bodies of research.(34) Remdesivir had its chemical origins in research on Tubercidin, a cytotoxic nucleoside produced by *Streptomyces tubercidicus* known to inhibit RNA polymerases. Foundational studies of the effects of 1’ modifications in the ribose of the C-nucleoside analog of Tubercidin demonstrated that nitrile substitution at this position produced a compound with low micro-molar activity against multiple viral strains including Parainfluenza 3 and SARS-CoV.(35) This compound, however, had unsuitable pharmaceutical properties, and required further modifications using established prodrug technologies.(32) One of the resulting compounds, GS-5734 (remdesivir), was shown to inhibit RdRp from HCV and had activity against Ebola virus.(36-38) Remdesivir was tested previously in clinical trials for Ebola, and while it was shown to be relatively safe, it proved to be inferior to other therapies.(39)

With the discovery and sequencing of COVID-19 in late 2019 and early 2020, it was recognized that the structure of the COVID-19 RdRp protein preserved essential features of the homologous nucleotide binding site of RdRp from SARS and MERS, suggesting that it could also be an inhibitor of COVID-19 RdRp.(40) *In vitro* studies confirmed that remdesivir was active against COVID-19,(41) and the first clinical trials began in February 2020. These trials provided sufficient evidence that remdesivir shortened the clinical course of the disease for Emergency Use Authorization at the end of April 2020.(42)

The quantitative analysis of research advance and maturation used in this study does not identify discrete milestones in the discovery or development of remdesivir. Rather, this approach posits that the broad corpus of published research—including research representing new discoveries, research refining details of biological structures or functions, research relating failed experiments, and research replicating previous findings—all contribute to the foundation of knowledge necessary for successful drug development. In the case of remdesivir, successful development required extensive prior knowledge of the structure-activity relationship of RdRp from different viral species as well as established strategies for circumventing the limitations of NcAn compounds as pharmaceutical products.

The relationship between the accumulation of research on RdRp and the emergence of effective RdRp inhibitors is consistent with previous studies demonstrating that few targeted therapeutics are approved before research on the biological target passes an established maturity point.(4, 5, 8) Specifically, while 97 products targeted to RdRp entered development since 1989, the first clinical successes were achieved only after research on both the drug target and parent drug structure passed the *established* point. The first approved nucleoside drugs (e.g., cytarabinine approved in 1969) were natural products discovered through phenotypic methods, which are commonly approved before there is a mature body of basic research on the drug target.(5, 8) Nevertheless, the current generation of nucleoside analogues designed to inhibit RdRp are also products of a mature body of research on nucleoside chemistry and pharmacology.

This analysis also demonstrates the substantial cost of establishing this mature body of foundational research. NIH-funding contributed to 20% of RdRp publications, totaling $1,875 million (2000–2019). Of this amount, $547 million was contributed before the *established* point in 2008. NIH funding also contributed to 21% of all NcAn publications, totaling $4,612 million (2000–2019). For both, the fraction of NIH-funded publications was lower than reported previously for other drug targets.(10, 11) This is consistent with the long-term decline in U.S. funding for university research,(43) but also associated with a sharp drop-off in the number of NIH-funded PMID related to RdRp observed after 2012.

The present results present a distinctly different picture of the public sector investment underlying remdesivir than a recent KEI Briefing Note.(20) That report focused exclusively on identifying NIH funding acknowledged in publications describing the development of remdesivir, as opposed to the basic research that made the design and discovery of this entity possible. The substantially higher estimate of the NIH contribution to remdesivir detailed in the present study is consistent with the observation that >90% of NIH funding contributing to drugs approved from 2010-2019 represented basic research on drug targets, rather than the drugs themselves (10,11), and the fact that the biopharmaceutical industry is primarily responsible for funding drug development.(6, 44-46) While case studies show that drug development programs in industry often benefit from public sector input,(45, 46), <10% of new drugs are first patented by academic institutions(47) and only half of drug patents cite prior art from public institutions.(48) It is worth noting that since COVID-19 was declared a public health emergency in January 2020, the federal government has assumed a more aggressive role in funding development of remdesivir and other therapies for COVID-19.

It is notable that while the large majority of the NIH-funded Projects Years related to RdRp and NcAn were investigator-initiated Research Projects (primarily R01), these grants represent only a small fraction of total Project Costs, less than the Costs associated with either Research Program Projects and Centers or Cooperative Agreements. This diversity of funding mechanisms reflects the broad contribution of NIH funding to investigator-initiated initiatives, research infrastructure, and mission-driven health initiatives as environed by the NIH Roadmap.(49)

There are several limitations to the present analysis. First, PubMed searches may fail to identify relevant publications describing enabling technologies or research performed before widely-recognized vocabularies emerge. Second, it is not possible to associate costs with every NIH-funded publication due to differences between the publication dates and Project Years. The method used here corrects for the estimated 3-year lag between publication dates and associated funding in RePORTER(50) by assigning costs for the final Project Year of each project to publications 1–4 years after the Project ended.(10) With this correction, the method associates costs with 90–91% of NIH-funded PMID and 79–80% of Project Years for RdRp and NcAn respectively. Third, this method does not include funding for research not reported through NIH RePORTER, such as funding from Department of Defense, philanthropies, and international sources. Thus, the total public sector funding for research underlying remdesivir is likely substantially greater than the amount described here. Finally, this analysis does not account for investments in applied research and development by industry, which are estimated to be as much as $1.5 billion in out-of-pocket costs for each new drug approval.(6)

These results demonstrate the critical role played by an existing foundation of basic research in the ability to rapidly develop remdesivir in response to the COVID-19 pandemic. In doing so, it argues for a federal commitment to sustained funding for a broad base of basic research, even when no imminent threat exists. This work also demonstrates the scale of the government and public sector investment in the research that enabled development of remdesivir and other RdRp inhibitors that are likely to be developed for treating COVID-19. This investment, along with that of industry, needs to be counted in developing policies to ensure the availability and affordability of the drugs that result from this research. As stated in an April 30, 2020 letter from Representatives Lloyd Doggett and Rosa DeLauro to HHS secretary Alex Azar “The substantial taxpayer investments in COVID-19 pharmaceutical research must be recognized.”(51)

## METHODS

### Data sources and analytical methods

Searches were performed in PubMed (www.pubmed.gov; accessed May 12, 2020) using the updated Automatic Term Mapping (May 2020) and search terms for remdesivir, RdRp, and NcAn described in Supplemental Table 1. Search terms were optimized by examining publication titles, abstracts, and descriptions of search results for randomly selected result pages and eliminating query terms that introduced irrelevant results. Publications were identified by PubMed Identifier (PMID) and publication year.

Methods for quantifying the contribution of NIH funding to publications identified in PubMed using NIH RePORTER (https://exporter.nih.gov/ExPORTER_Catalog.aspx; accessed May 5, 2020), have been described previously.(10) Briefly, PMID were associated with fiscal years of program funding (Program Years) from 1980–2019 using the “Link Table,” and annual Project Costs from 2000–2019 identified using the “Projects Table.” Publications occurring before the first year of the grant award or more than four years after the last year of the grant were excluded. Publications 1–4 years after the last year of the grant were associated with the Project Costs of the last year. PMID related to remdesivir are included with data for the target RdRp. Grant categories were derived from NIH Types of Grant Programs 2020 (https://grants.nih.gov/grants/funding/funding_program.htm; accessed May 2020). Costs are given in constant dollars inflation adjusted to 2018 using the U.S. Bureau of Labor Statistics All Urban Consumer Prices (Current Series) (https://www.bls.gov/data/; accessed May, 2020).

Text analysis was performed on abstracts, titles, MeSH descriptors, and MeSH qualifiers to quantify mentions of specific positive-strand ssRNA or negative-strand ssRNA viruses targeted in drug discovery programs(21) as well as terms related to oncology and immunology. All analyses were performed in PostgreSQL, Excel and Tableau.

### Technology Maturation Model

The bibliographic Technology Innovation Maturation Evaluation (TIME) model fits the cumulative number of publications resulting from a PubMed search to an exponentiated logistic function as described previously.(4) The equation has the form:

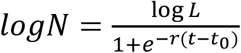

Where *N* is the number of publications, *L* is the presumed upper limit of publications, *r* is the growth rate, *t* is time, and *t*_0_ is midpoint of the exponential growth. This asymmetric sigmoidal function, exhibits the common logistic sigmoid function over log scales. The parameters were fit to time series publication data using a non-linear least squares implementation of the Levenberg-Marquardt algorithm (LMFIT, version 1.0.1).

The *initiation* and *established* points, representing the beginning and end of the exponential growth phase are defined as the points of maximum and minimum acceleration respectively, or log*N*’’(*t*)_max,min_. These points are analytically determined by:

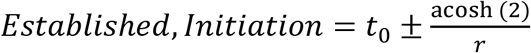

## Data Availability

All data is freely available. All code is posted in GitHub.

https://github.com/

## Author contributions

E.G.C., M.J.J., Z.F.W., and F.D.L. designed research; E.G.C., M.J.J., Z.F.W., and F.D.L. performed research; E.G.C., M.J.J., Z.F.W., and F.D.L. analyzed data; and E.G.C., M.J.J., Z.F.W., and F.D.L. wrote the paper; F.D.L. supervised the project and acquired financial support.

## ACKNOWLEDGEMENTS

This work was supported by grants from the National Biomedical Research Foundation and the Institute for New Economic Thinking to Bentley University. Abbreviations: RNA-dependent RNA polymerase, RdRp; Nucleoside analogue, NcAn; PubMed Identifier, PMID; Technology Innovation Maturation Evaluation model, (TIME model). The authors thank Drs. Michael Boss, Franklin Bright, and Nancy Hsiung as well as Ethan Benton for their contributions to this research.

## REFERENCES

1. FDA (2020) Emergency Use Authorization letter for remdesivir, dated May, 1, 2020. (Food and Drug Administration).

2. Z.S. Morris, S. Wooding, J. Grant, The answer is 17 years, what is the question: understanding time lags in translational research. Journal of the Royal Society of Medicine 104(12):510–520 (2011).

3. J. Eder, R. Sedrani, C. Wiesmann, The discovery of first-in-class drugs: origins and evolution. Nature Reviews Drug Discovery 13(8):577–587 (2014).

4. L.M. McNamee, M.J. Walsh, F.D. Ledley, Timelines of translational science: from technology initiation to FDA approval. PLoS ONE 12(5):e0177371 (2017).

5. J.M. Beierlein et al., Landscape of innovation for cardiovascular pharmaceuticals: from basic science to new molecular entities. Clinical Therapeutics 39(7):1409–1425 (2017).

6. J.A. DiMasi, H.G. Grabowski, R.W. Hansen, Innovation in the pharmaceutical industry: new estimates of R&D costs. Journal of Health Economics 47:20–33 (2016).

7. PCAST, Report to the President on Propelling Innovation in Drug Discovery, Development, and Evaluation (2012).

8. L.M. McNamee, F.D. Ledley, Modeling timelines for translational science in cancer; the impact of technological maturation. PLoS ONE 12(3):e0174538 (2017).

9. H. Moses et al., The anatomy of medical research: US and international comparisons. JAMA 313(2):174–189 (2015).

10. E.G. Cleary, J.M. Beierlein, N.S. Khanuja, L.M. McNamee, F.D. Ledley, Contribution of NIH funding to new drug approvals 2010–2016. Proceedings of the National Academy of Sciences 115(10):2329–2334 (2018).

11. E.G. Cleary, M. Jackson, A. Acevedo, F.D. Ledley, Characterizing the public sector contribution to drug discovery and development: the role of government as a first investor. Institute for New Economic Thinking (2020).

12. E.G. Cleary, F.D. Ledley, NIH funding for research underlying new cancer therapies. The Lancet. Oncology 21(6):755–757 (2020).

13. C.M. Christensen, Exploring the limits of the technology S-curve. Part I: component technologies. Production and Operations Management 1(4):334–357, (1992).

14. R.N. Foster, Effective R&D operations in the’80s: boosting the payoff From R&D. Research Management 25(1):22–27 (1982).

15. J.M. Beierlein, L.M. McNamee, F.D. Ledley, As technologies for nucleotide therapeutics mature, products emerge. Molecular Therapy-Nucleic Acids 9:379–386 (2017).

16. M.D. Whittington, J.D. Campbell, Alternative pricing models for remdesivir and other potential treatments for COVID-19. (Institute for Clinical and Economic Review) (2020).

17. E. Silverman, Lawmakers ask HHS to ensure Gilead’s remdesivir is affordable if U.S. taxpayers funded R&D. STAT News (May 1, 2020..

18. A. Liu, Gilead should be allowed “real pricing” for remdesivir-and a sizable profit, analyst says. In Fierce Phama, (June 11, 2020..

19. G. Kolata, How remdesivir, new hope for Covid-19 patients, was resurrected. New York Times (May 1, 2020..

20. K. Ardizzone, Role of the federal government in the development of remdesivir. In KEI Briefing Note (2020).

21. E. De Clercq, G. Li, Approved antiviral drugs over the past 50 years. Clinical Microbiology Reviews 29(3):695–747 (2016).

22. O.L. Bryan-Marrugo et al., History and progress of antiviral drugs: from acyclovir to direct-acting antiviral agents (DAAs) for Hepatitis C. Medicina Universitaria 17(68):165–174 (2015).

23. C.M. Galmarini, J.R. Mackey, C. Dumontet, Nucleoside analogues and nucleobases in cancer treatment. The Lancet Oncology 3(7):415–424 (2002).

24. V.L. Damaraju et al., Nucleoside anticancer drugs: the role of nucleoside transporters in resistance to cancer chemotherapy. Oncogene 22(47):7524–7536 (2003).

25. J.D. Westbrook, S.K. Burley, How structural biologists and the protein data bank contributed to recent FDA new drug approvals. Structure 27(2):211–217 (2019).

26. V.J. Venditto, F.C. Szoka Jr., Cancer nanomedicines: so many papers and so few drugs! Advanced Drug Delivery Reviews. 65(1):80–88 (2013).

27. D. Baltimore, R.M. Franklin, A new ribonucleic acid polymerase appearing after mengovirus infection of L-cells. J Biol Chem 238:3395–3400 (1963).

28. D. Baltimore, R.M. Franklin, J. Callender, Mengovirus-induced inhibition of host ribonucleic acid and protein synthesis. Biochimica et Biophysica Acta (BBA)-Specialized Section on Nucleic Acids and Related Subjects 76:425–430 (1963).

29. E.H. Simo, Evidence for the nonparticipation of DNA in viral RNA synthesis. Virology 13(1):105–118 (1961).

30. S. Venkataraman, B.V. Prasad, R. Selvarajan, RNA dependent RNA polymerases: insights from structure, function and evolution. Viruses 10(2):76 (2018).

31. A. Yssel, J. Vanderleyden, H. Steenackers, Repurposing of nucleoside-and nucleobase-derivative drugs as antibiotics and biofilm inhibitors. Journal of Antimicrobial Chemotherapy 72(8):2156–2170 (2017).

32. Y. Mehellou, H.S. Rattan, J. Balzarini, The ProTide Prodrug Technology: From the Concept to the Clinic: Miniperspective. Journal of Medicinal Chemistry 61(6):2211–2226 (2017).

33. L.W. Peterson, C.E. McKenna, Prodrug approaches to improving the oral absorption of antiviral nucleotide analogues. Expert Opinion on Drug Delivery 6(4):405–420 (2009).

34. R.T. Eastman et al., Remdesivir: a review of its discovery and development leading to emergency use authorization for treatment of COVID-19. ACS Central Science (2020).

35. A. Cho et al., Synthesis and antiviral activity of a series of 1′-substituted 4-aza-7, 9-dideazaadenosine C-nucleosides. Bioorganic & Medicinal Chemistry Letters 22(8):2705–2707(2012).

36. C.J. Gordon, E.P. Tchesnokov, J.Y. Feng, D.P. Porter, M. Götte, The antiviral compound remdesivir potently inhibits RNA-dependent RNA polymerase from Middle East respiratory syndrome coronavirus. Journal of Biological Chemistry 295(15):4773–4779 (2020).

37. A.J. Brown et al., Broad spectrum antiviral remdesivir inhibits human endemic and zoonotic deltacoronaviruses with a highly divergent RNA dependent RNA polymerase. Antiviral Research 169:104541 (2019).

38. D. Siegel et al., Discovery and Synthesis of a Phosphoramidate Prodrug of a Pyrrolo [2, 1-f][triazin-4-amino] Adenine C-Nucleoside (GS-5734) for the Treatment of Ebola and Emerging Viruses. (ACS Publications) (2017).

39. S. Mulangu et al., A randomized, controlled trial of Ebola virus disease therapeutics. New England Journal of Medicine 381(24):2293–2303 (2019).

40. W-C. Ko et al., Arguments in favour of remdesivir for treating SARS-CoV-2 infections. International Journal of Antimicrobial Agents (2020).

41. M. Wang et al., Remdesivir and chloroquine effectively inhibit the recently emerged novel coronavirus (2019-nCoV) in vitro. Cell Research 30(3):269–271 (2020).

42. Y. Wang et al., Remdesivir in adults with severe COVID-19: a randomised, double-blind, placebo-controlled, multicentre trial. The Lancet 395(10236):1569–1578 (2020).

43. R.D. Atkinson, C. Foote, US funding for university research continues to slide. (Information Technology and Innovation Foundation) (2019).

44. O.J. Wouters, M. McKee, J. Luyten, Estimated research and development investment needed to bring a new medicine to market, 2009.2018. Jama 323(9):844–853 (2020).

45. R. Chakravarthy, K. Cotter, J. DiMasi, C-P. Milne, N. Wendel, Public-and private-sector contributions to the research and development of the most transformational drugs in the past 25 years: from theory to therapy. Therapeutic Innovation & Regulatory Science 50(6):759–768 (2016).

46. B. Zycher, J.A. DiMasi, C-P. Milne, Private sector contributions to pharmaceutical science: thirty-five summary case histories. American Journal of Therapeutics 17(1):101–120 (2010).

47. A.J. Stevens et al., The role of public-sector research in the discovery of drugs and vaccines. New England Journal of Medicine 364(6):535–541 (2011).

48. B.N. Sampat, F.R. Lichtenberg, What are the respective roles of the public and private sectors in pharmaceutical innovation? Health Affairs 30(2):332–339 (2011).

49. E. Zerhouni, The NIH roadmap. Science 203(5642):63–72.

50. K.W. Boyack, P. Jordan, Metrics associated with NIH funding: a high-level view. Journal of the American Medical Informatics Association 18(4):423–431 (2011).

51. L. Dogett, R. DeLauro, Letter to HHS Secretary Alex Azar, April 30, 2020.

